# The facilitators and barriers to improving functional activity and wellbeing in people with dementia: A qualitative study from the Process Evaluation of Promoting Activity, Independence and Stability in Early Dementia

**DOI:** 10.1101/2022.12.20.22283555

**Authors:** Claudio Di Lorito, Veronika van der Wardt, Kristian Pollock, Louise Howe, Vicky Booth, Pip Logan, John Gladman, Tahir Masud, Roshan das Nair, Sarah Goldberg, Kavita Vedhara, Rebecca O’Brien, Emma Adams, Alison Cowley, Alessandro Bosco, Jennie Hancox, Clare Burgon, Rupinder Bajwa, Juliette Lock, Annabelle Long, Maureen Godfrey, Marianne Dunlop, Rowan H. Harwood

## Abstract

**Background:** The PRomoting Activity, Independence and Stability in Early Dementia (PrAISED) study delivered an exercise and functional activity programme to participants living with dementia. A Randomised Controlled Trial (RCT) showed no measurable benefits in activities of daily living, physical activity or quality of life.

**Objective:** To explore participants’ responses to PrAISED and explain the mechanisms behind a complex intervention that did not lead to expected health gains.

**Methods:** A process evaluation using qualitative methods, comprising interviews and researcher notes

**Setting:** Data were collected in participants’ homes or remotely by telephone or videoconferencing.

**Sample:** Eighty-eight interviews were conducted with 44 participants living with dementia (n = 32 intervention group; n = 12 control group) and 39 caregivers. Sixty-nine interviews were conducted with 26 therapists.

**Results:** Participants valued the intervention as proactively addressing health issues that were of concern to them, and as sources of social contact, interaction, information, and advice. Facilitators to achieving positive outcomes included perceiving progress toward desired goals, positive expectations, therapists’ skills and rapport with participants, and caregiver support. Barriers included: cognitive impairment, which prevented independent engagement and carryover between sessions; chronic physical health problems and intercurrent acute illness and injury; ‘tapering’ (progressively infrequent supervision intended to help develop habits and independent activity); and the COVID-19 pandemic.

**Conclusions:** Interventions aiming to maintain activity, independence and stability may not be appropriate in the context of dementia even in the mild stages of the condition. Various factors affected outcomes including caregiver support, rapport with therapists, availability of supervision, motivational factors, and the limitations of remote delivery. The effects of cognitive impairment, multimorbidity and frailty overwhelmed any positive impact of the intervention. Maintenance of functional ability is valued, but in the face of inevitable progression of disease, other less tangible outcomes become important, challenging how we frame ‘health gain’ and trial outcomes.

## Introduction

Dementia is a neurodegenerative condition characterised by a progressive deterioration of both cognitive and motor functioning leading to a loss of independence, reduced quality of life and increased risk of injuries and hospitalisation [1-8]. A number of functional activity, physical activity and exercise intervention programmes have been developed to help people living with dementia to maintain their independence for longer [9,10], including the Promoting Activity, Independence and Stability in Early Dementia (PrAISED) intervention [11,12].

We evaluated the PrAISED intervention in a Randomised Controlled Trial (RCT), randomising 365 participants across 5 sites in England to an intervention arm or a control group (brief falls assessment and advice) [12]. Participants in the intervention arm took part in an individually tailored programme comprising physical exercises (i.e., progressive strength, balance, and dual task); functional activities (activities of daily living, ADL, with an element of physical activity, such as going shopping); promotion of inclusion in community life (e.g., signposting physical exercise group classes); risk enablement (positive risk-taking); and environmental assessment (accessibility and safety issues at home). They received up to 50 home therapy sessions over 12-months from a multidisciplinary team comprising physiotherapists (PTs), occupational therapists (OTs) and rehabilitation support workers (RSWs) (n = 68) [12]. The sessions were intended to teach and supervise exercise and functional activities, monitor progress, and adjust the programme. Participants were asked to undertake exercise activities between therapy sessions.

PrAISED delivered 5,356 therapy visits between October 2018 and June 2022. Participants recorded exercise on a monthly calendar and reported undertaking a mean of 121 minutes/week of PrAISED exercise. The RCT showed no measurable benefits in ADL, increased physical activity, quality of life or any other of the battery of health status measures studied [13].

PrAISED was a complex intervention because of its many interacting components (functional and physical exercises), the number of agents involved (people living with dementia, caregivers, therapists) and the different contexts (social and cultural) within which the programme was implemented [14]. When evaluating complex interventions, process evaluations are essential complements to RCTs [14]. A process evaluation identifies mechanisms of impact: (participant-level factors that affect the emergence of outcomes) and contextual factors (characteristics of the physical, cultural and social contexts) that affect the emergence of outcomes.

The aims of the PrAISED process evaluation were to investigate participants’ responses to the programme, and to explain the mechanisms behind a complex intervention that did not lead to expected health gains.

## Methods

This study followed Medical Research Council (MRC) guidance for process evaluation [14]. The protocols have been published [15,16]. We report on implementation in terms of reach, dose, fidelity, and adaptation elsewhere [13]. This study adopted a qualitative design, based on interviews, and researchers’ notes.

### Sample

Participants living with dementia and their caregivers were purposively selected to obtain a diverse sample in relation to age, gender, ethnicity, location, relationship status, and living status. Participants from the intervention and control groups were selected for comparison purposes.

Therapists were recruited based on their availability. Sample size was based on conceptual density (i.e., gathering data until a *sufficient depth* of understanding was reached) [17], which was agreed upon by two researchers (CDL and VvdW).

### Data collection

Semi-structured interviews were conducted with participants living with dementia and their caregivers (as dyads or individually, depending on preference), and with therapists (individually).

Two sets of interviews took place in the participants’ homes and in the therapists’ offices (Appendix 1): pre and during the COVID-19 pandemic. For the first set, the interviews took place at month six and month 12 of participants’ involvement in PrAISED. Interview topic guides (Appendices 2 and 3) were co-developed with two Patient and Public Involvement (PPI) co-researchers with lived experience of caring for someone living with dementia (MG and MD) to ensure that the interview schedule was relevant, meaningful, and accessible. The topic guide was informed by the Physical Activity Behaviour change Theoretical model in dementia (PHYT-in-dementia) [18,19], which identified a set of factors mediating intervention experience/outcomes, e.g., autonomy/independence, motivation. For each factor, several prompts were developed, but a flexible approach was adopted to explore emerging themes. All interviews were conducted by the first author, and eight interviews with participants and caregivers were co-facilitated with one PPI co-researcher. Written consent from all interviewees was obtained prior to the interview.

The second set of interviews, taking place following COVID-19 restrictions in England (March 2020), aimed to monitor the impact of restrictions on the delivery and reception of PrAISED. The original interview topic guides were adapted (Appendices 4 and 5). Interviews were conducted by telephone or video call and were undertaken by CDL. Verbal consent from all interviewees was audio recorded.

### Data analysis

All interviews were audio recorded, transcribed verbatim and anonymised. Participant/caregiver/therapist IDs were changed from the original study for anonymisation and they are not known to anyone outside the research group. Transcripts were analysed through inductive thematic analysis (20). CDL and two PPI co-researchers (MG and MD) examined the transcripts independently of each other and made annotations/reflections on emerging mechanisms of impact and contextual factors. The independent coders convened to discuss their annotations and create a tentative list of mechanisms of impact and contextual factors. The list was passed to a fourth (VvdW) and fifth researcher (AB), who provided feedback, which was integrated to finalise a coding scheme. This featured an operational definition for each mechanism of impact and contextual factor, data to be coded within each (for replicability) and participants’ quotations (Table 1). Four co-authors (MG, MD, VvdW and KP) checked the final analysis.

**Table 1.**
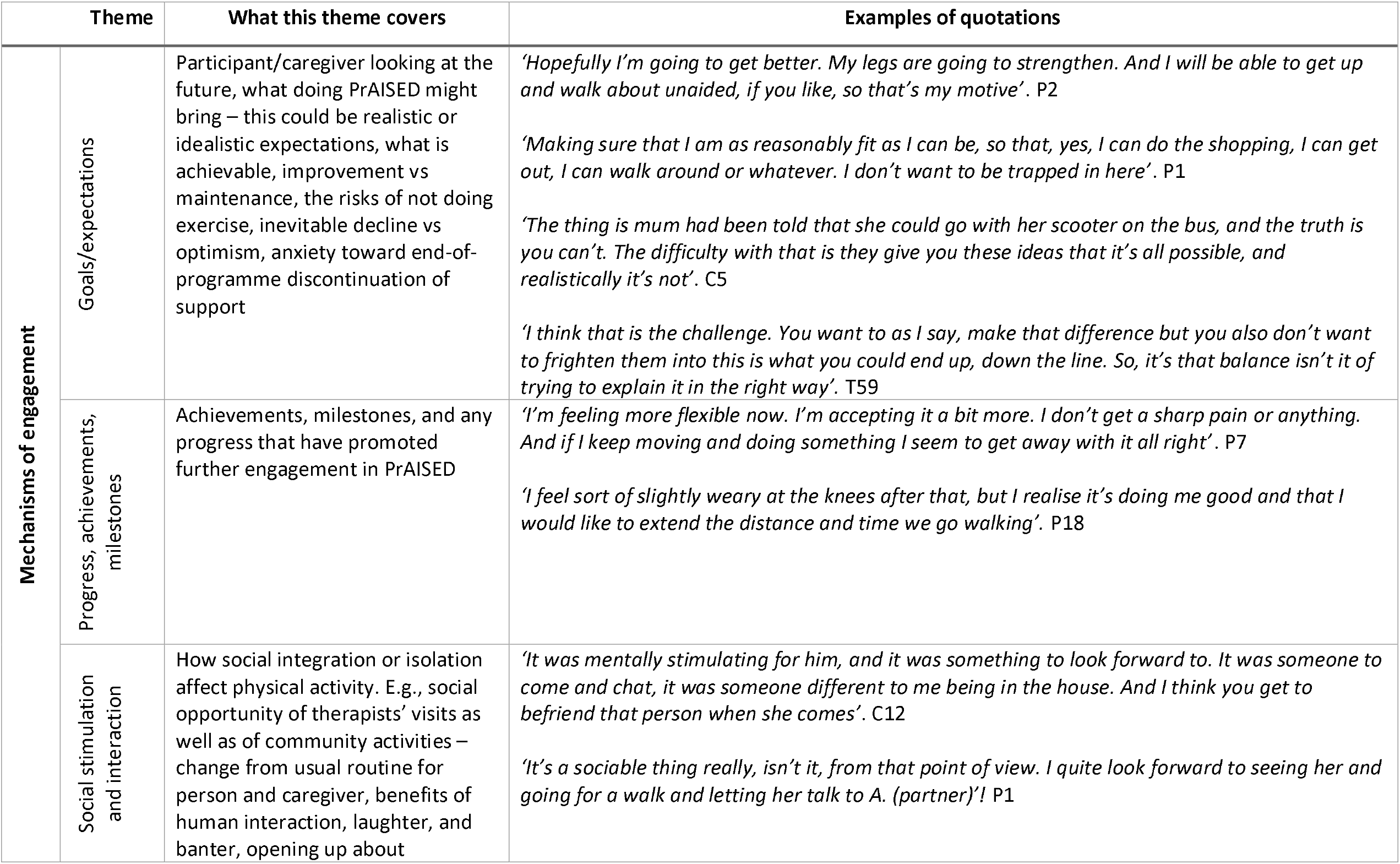
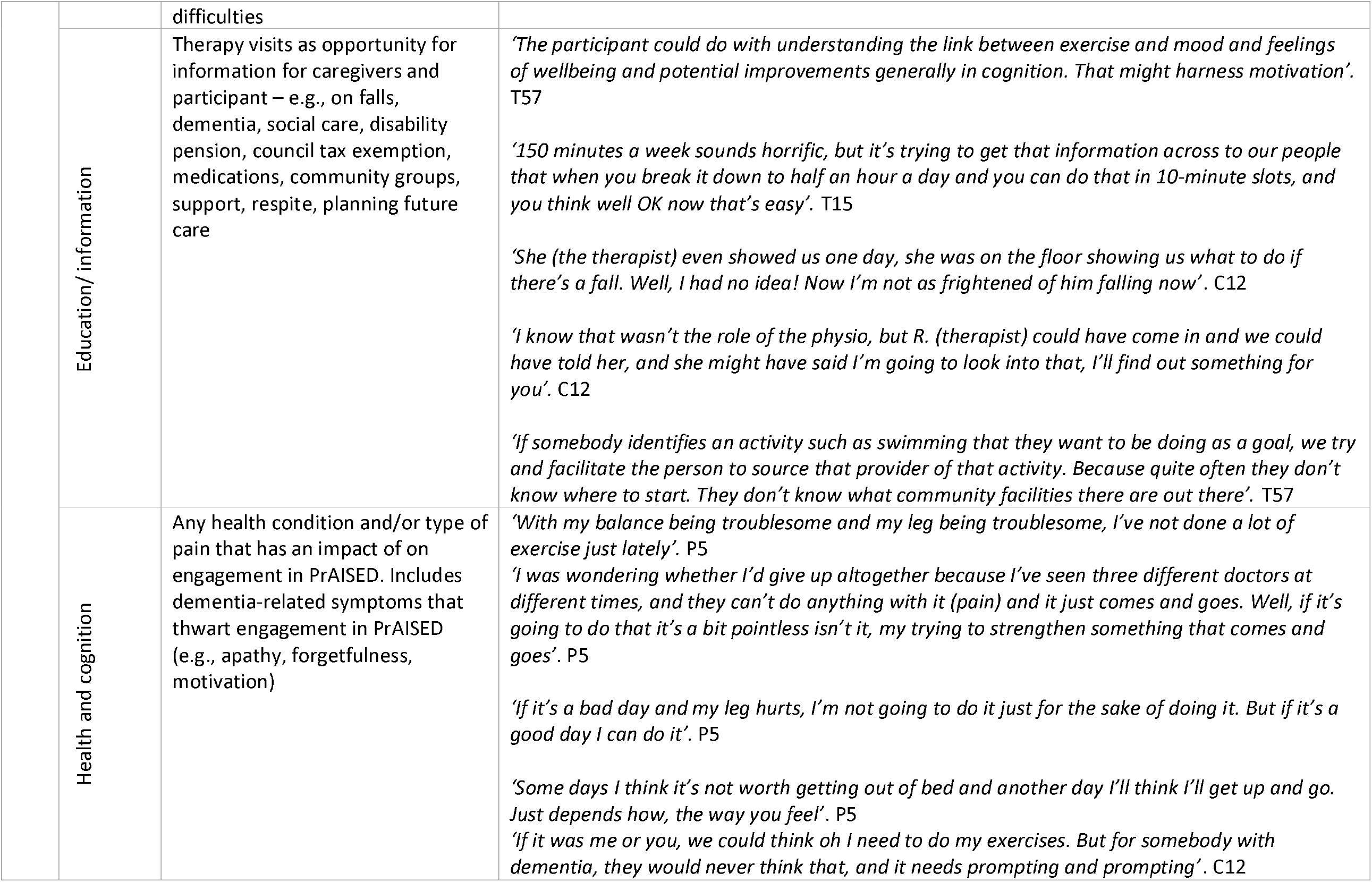
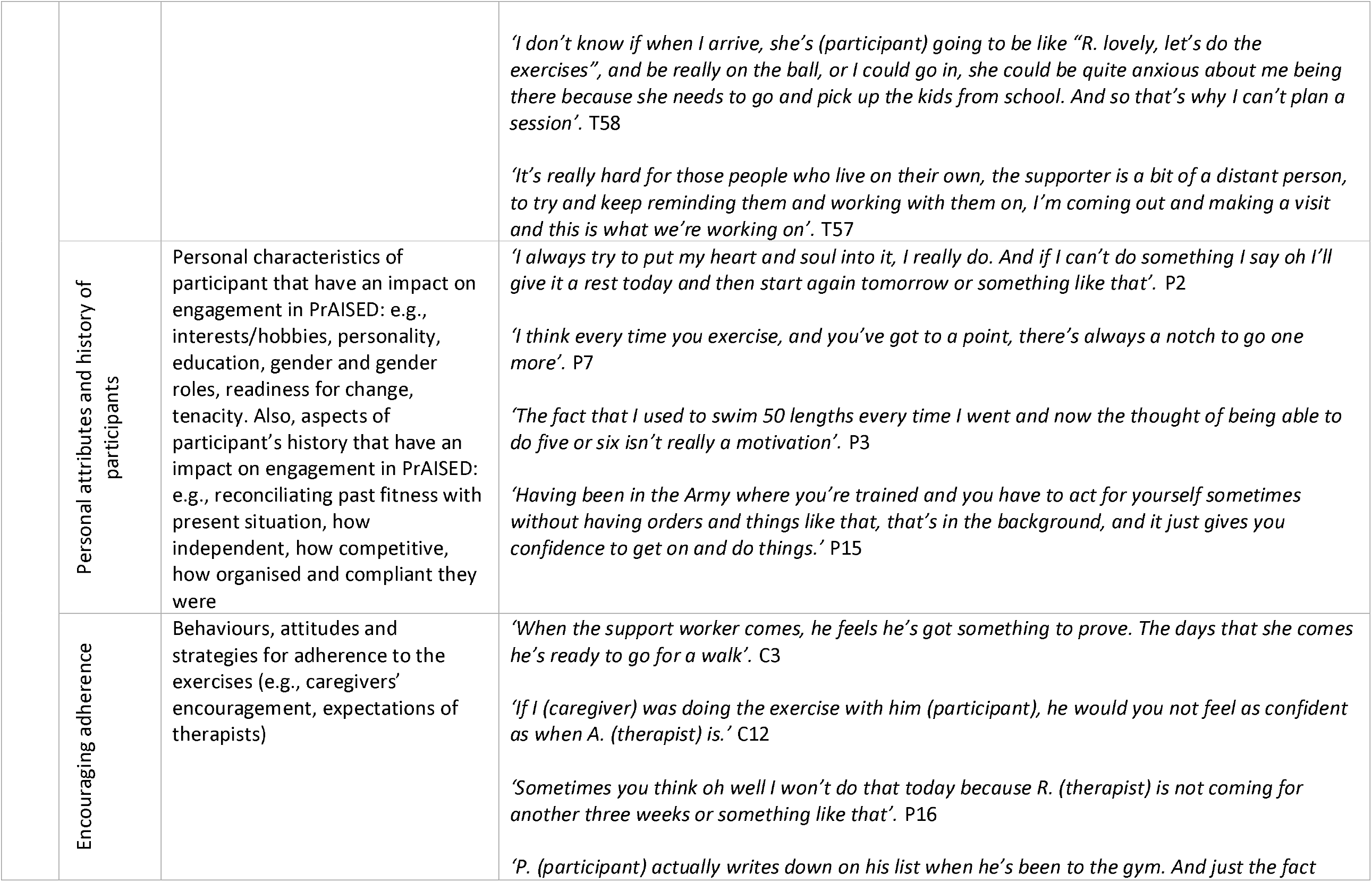
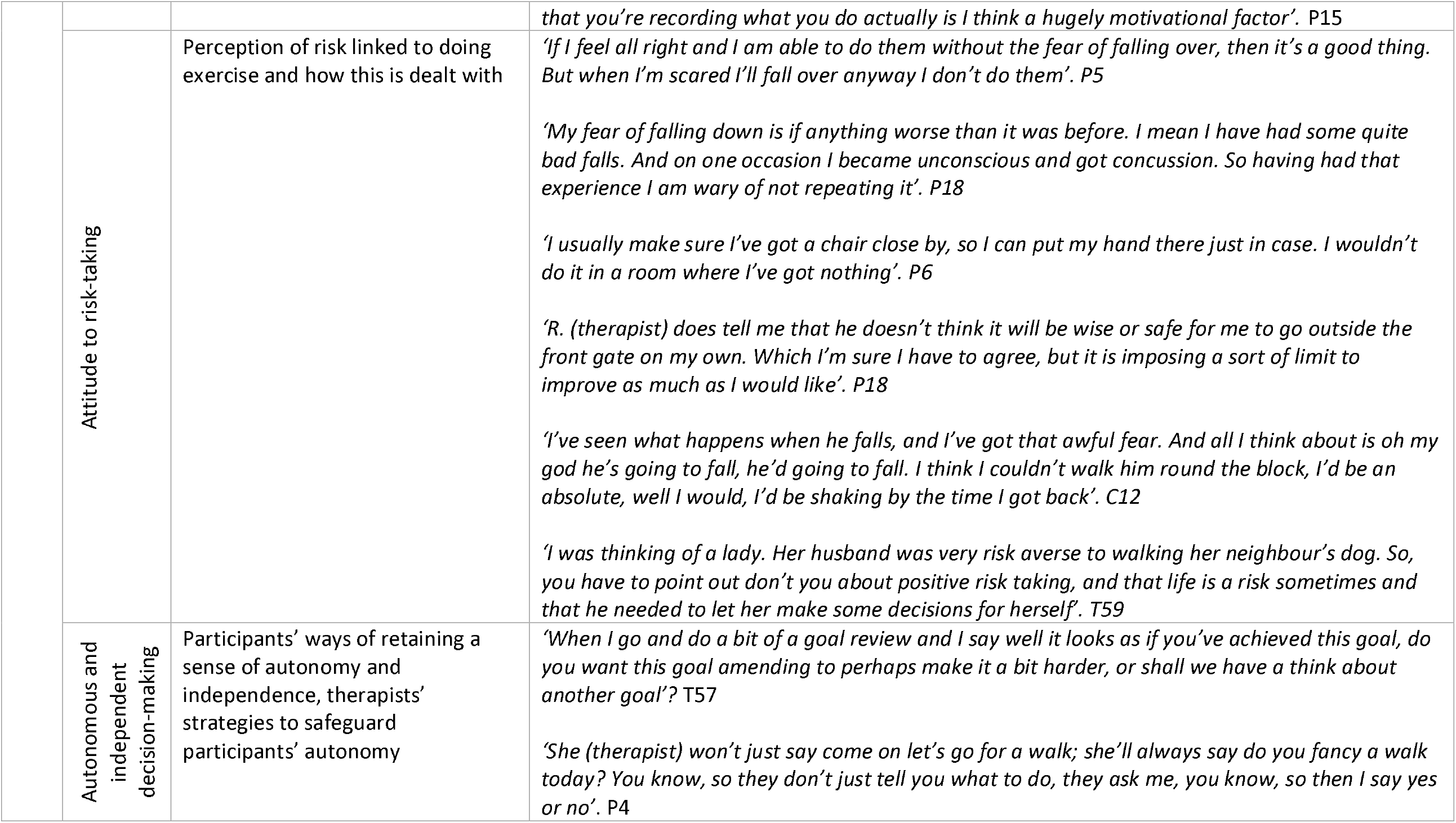
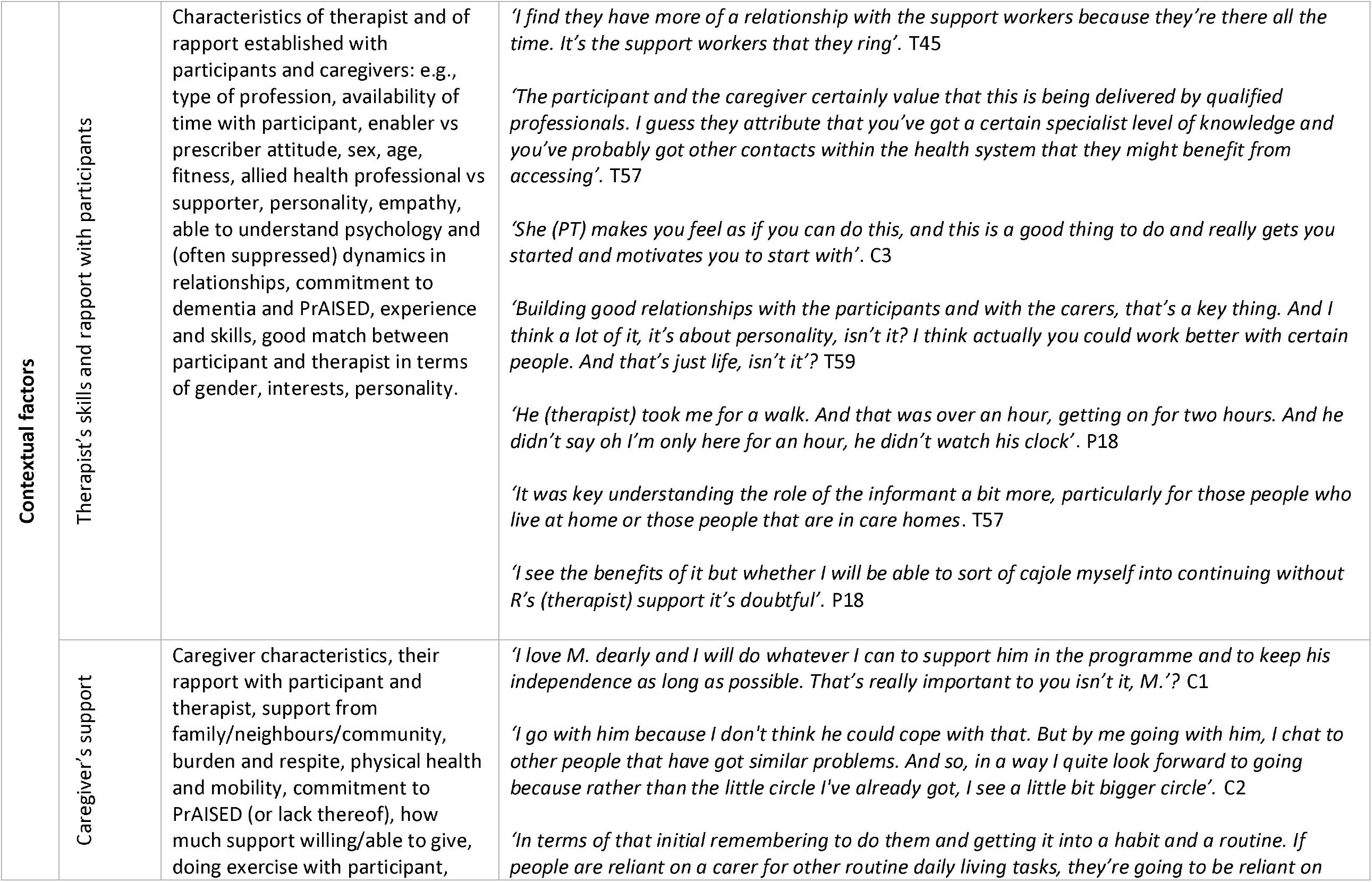
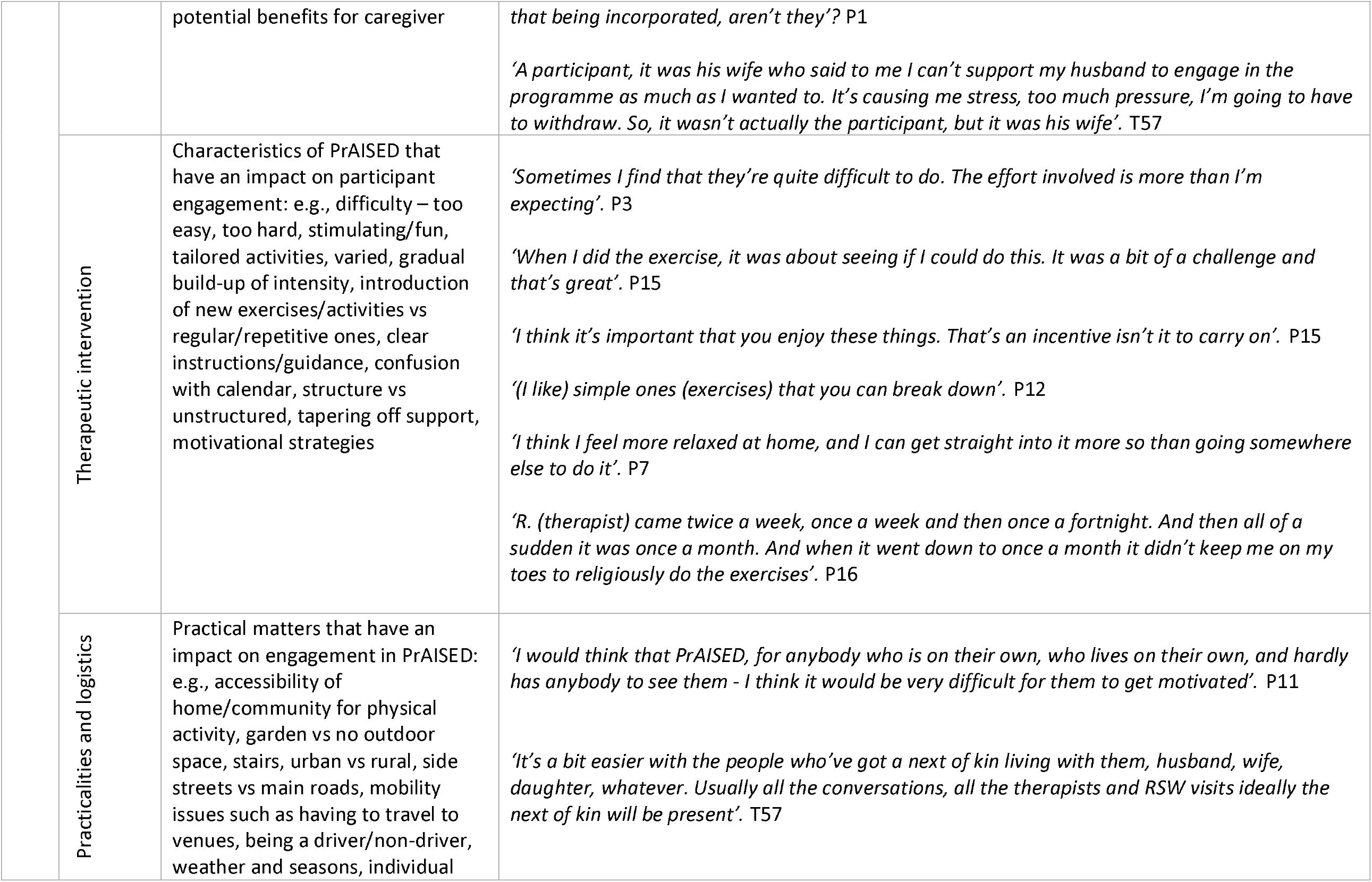
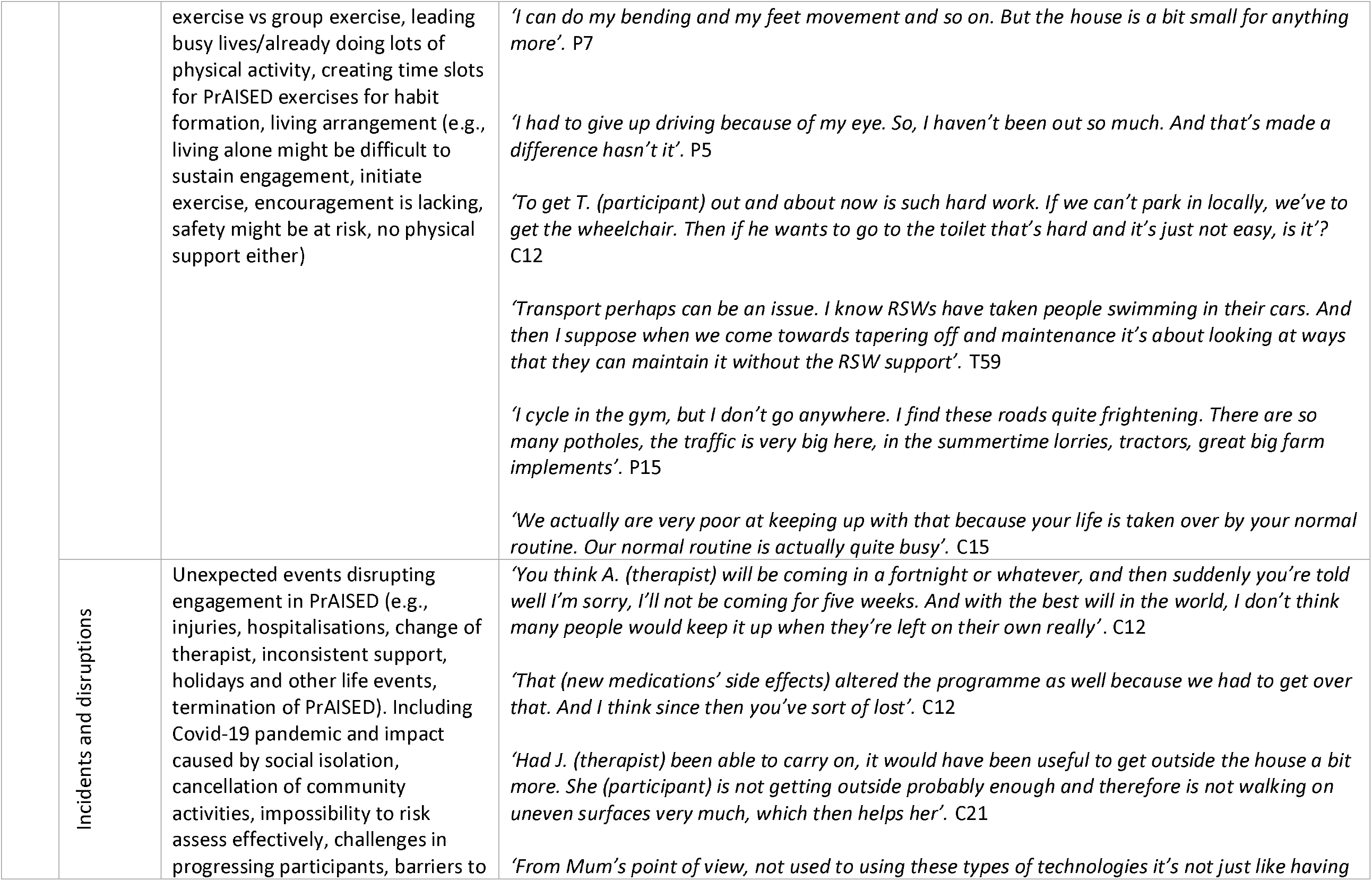
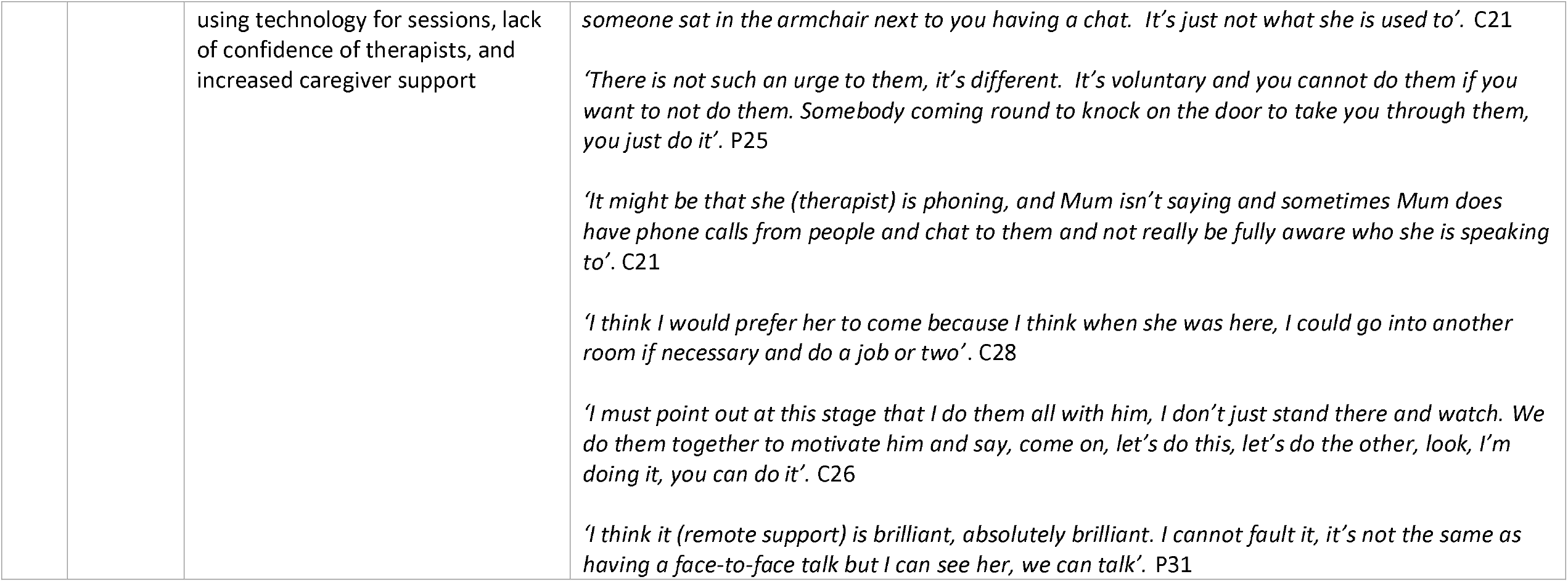
Mechanisms of engagement and contextual factors

Three co-authors (CDL, VvdW and RH) (independently of each other) included each identified mechanism of impact and contextual factor into one of the two categories of facilitators and barriers. The three authors then convened to agree on a definitive list. Based on interview data and personal reflections of the co-researchers (CDL, MG and MD) annotated after each interview session, four case-study vignettes (Appendix 6) and an ecological system model (Fig. 1) were produced to illustrate the complex interaction between barriers and facilitators in generating outcomes.

**Figure 1.**
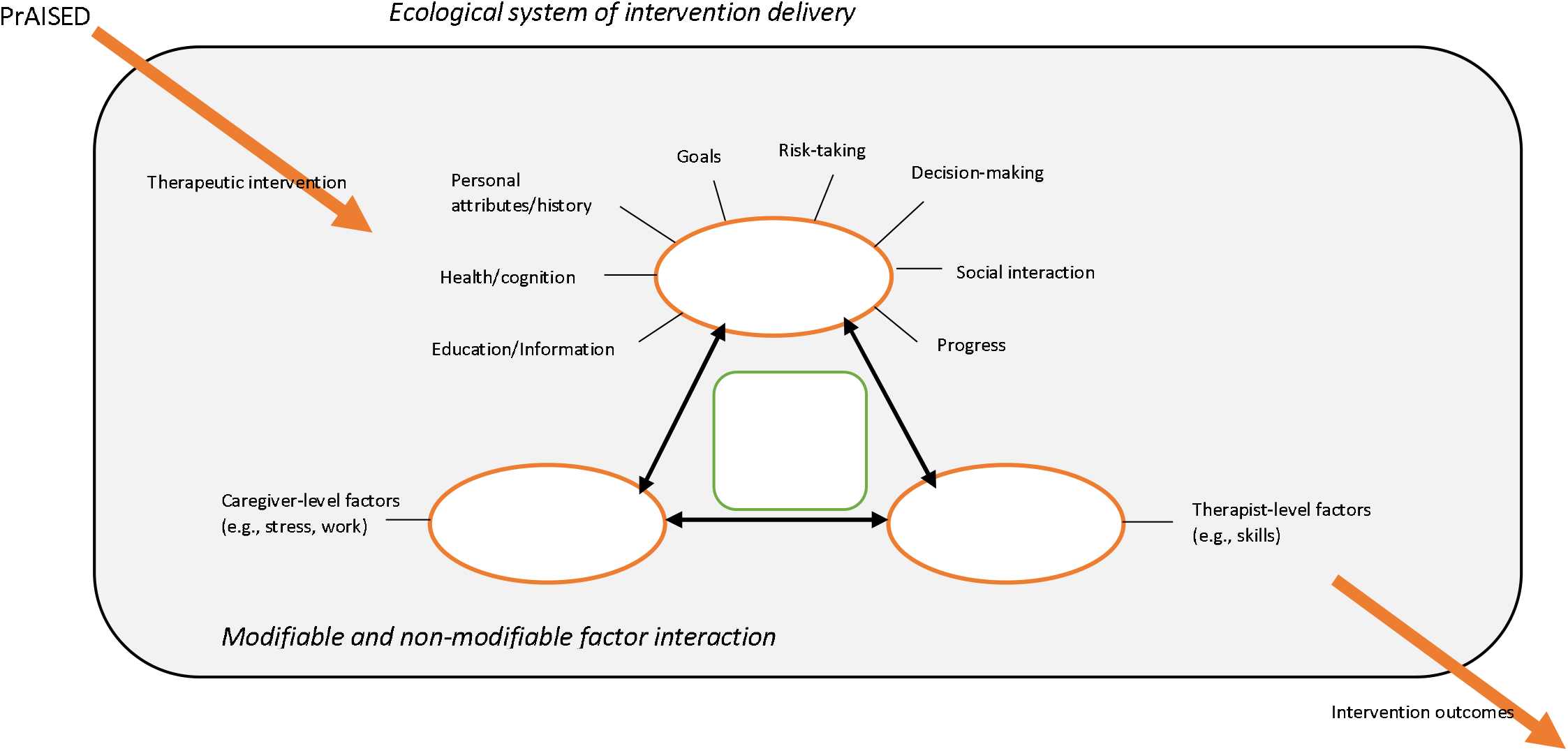
Ecological system model.

### Ethics

The PrAISED RCT and process evaluation received ethical approval from the Bradford-Leeds Research Committee (18/YH/0059).

## Results

Eighty-eight interviews were conducted with 44 participants living with Mild Cognitive Impairment (MCI) or dementia (mean age: 79 years, 95% white ethnicity, 84% living alone, n = 12 control and n = 32 intervention group) and 39 caregivers (mean age: 72 years). The interviews were dyadic (i.e., participant living with dementia and caregiver) face-to-face (n = 40), dyadic on the phone (n = 36), individual (i.e., participant living with dementia only) on the telephone (n = 10) and dyadic on webcam (n = 2). Sixty-nine interviews were conducted with 26 therapists (n = 8 physiotherapists, n = 8 occupational therapists, n = 10 rehabilitation support workers; Table 2).

**Table 2.**
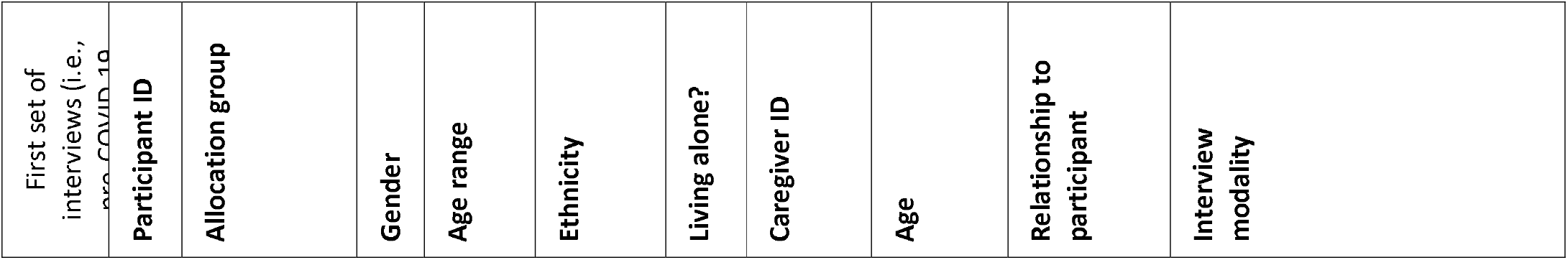
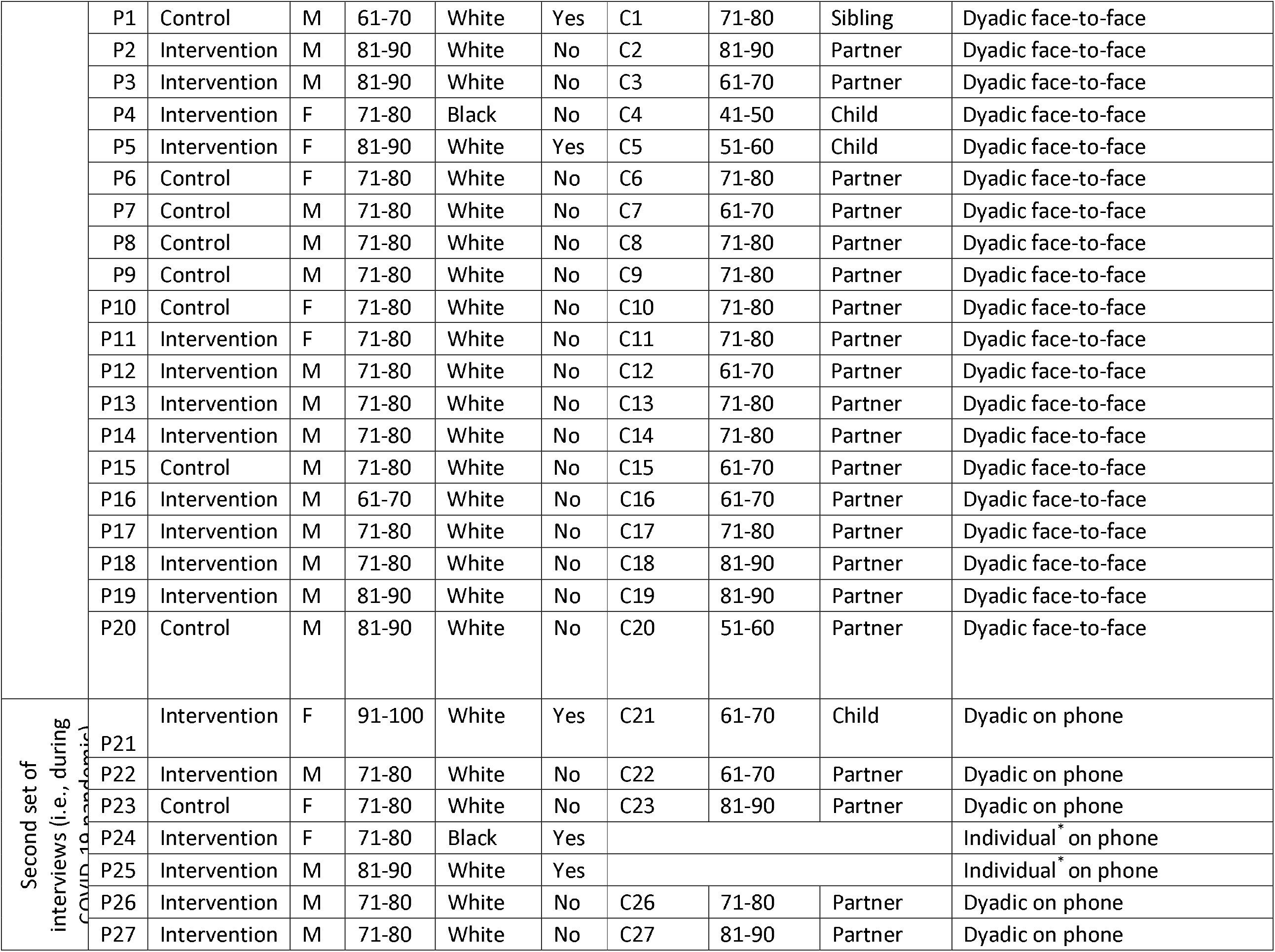
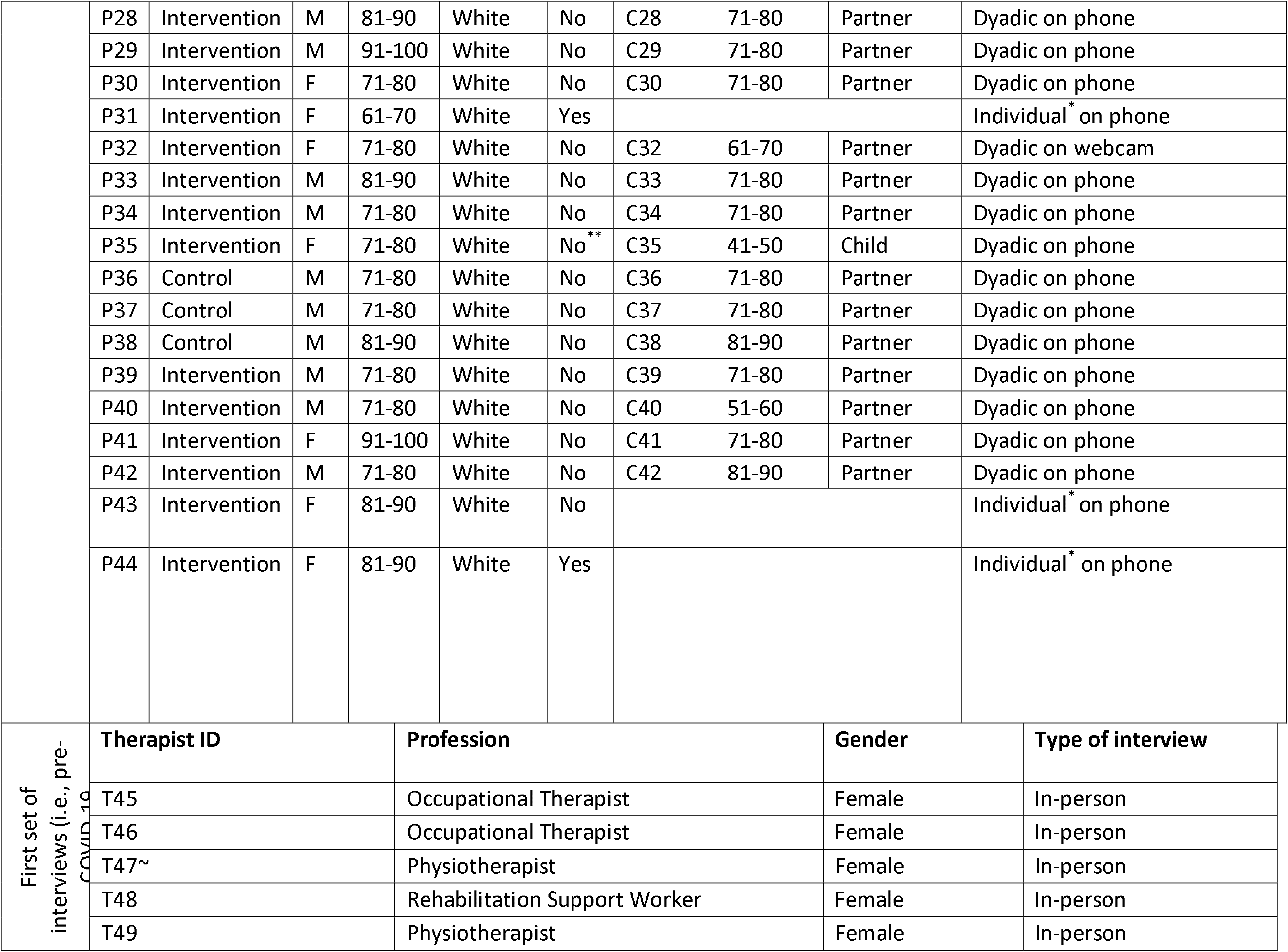
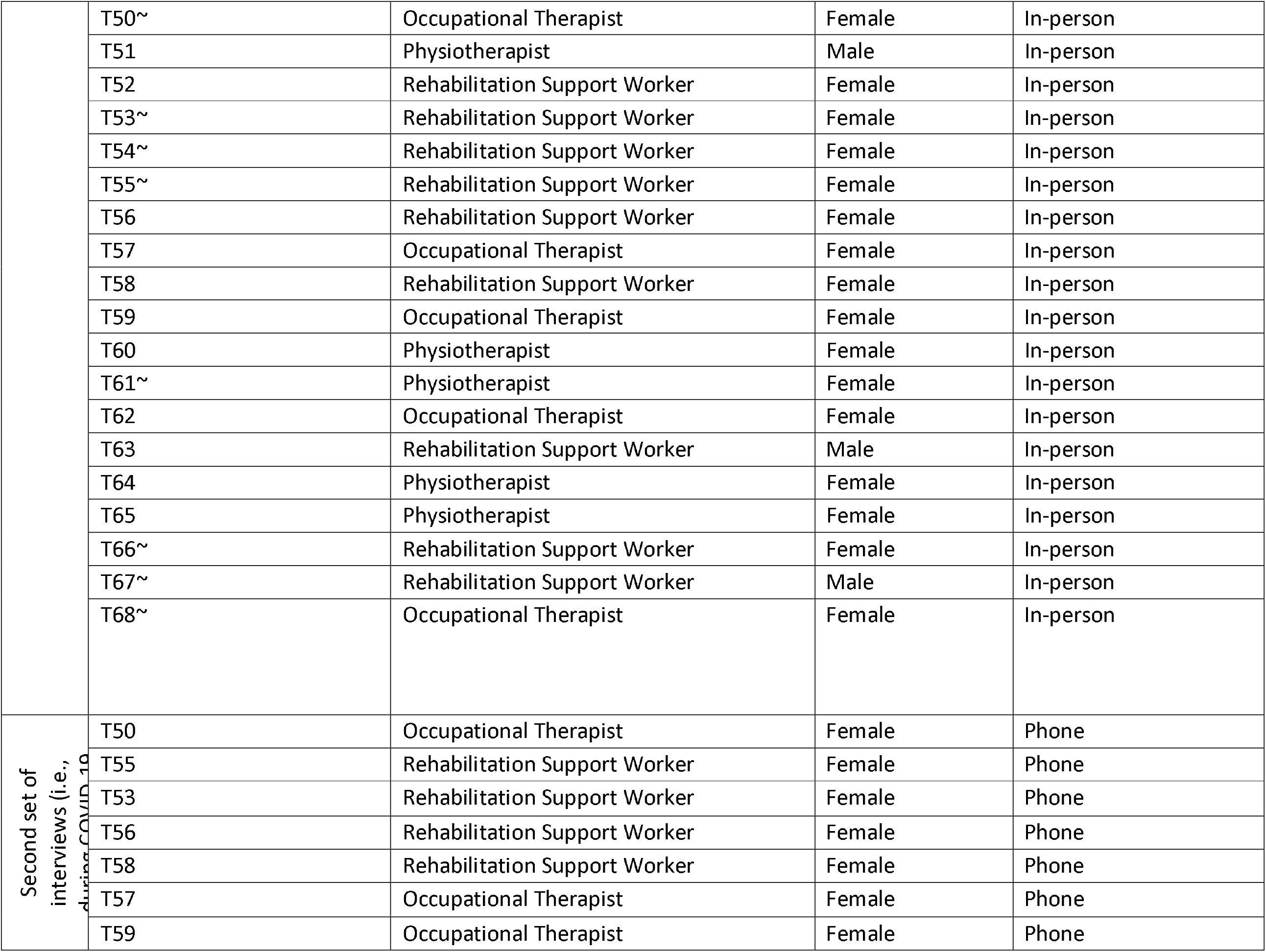
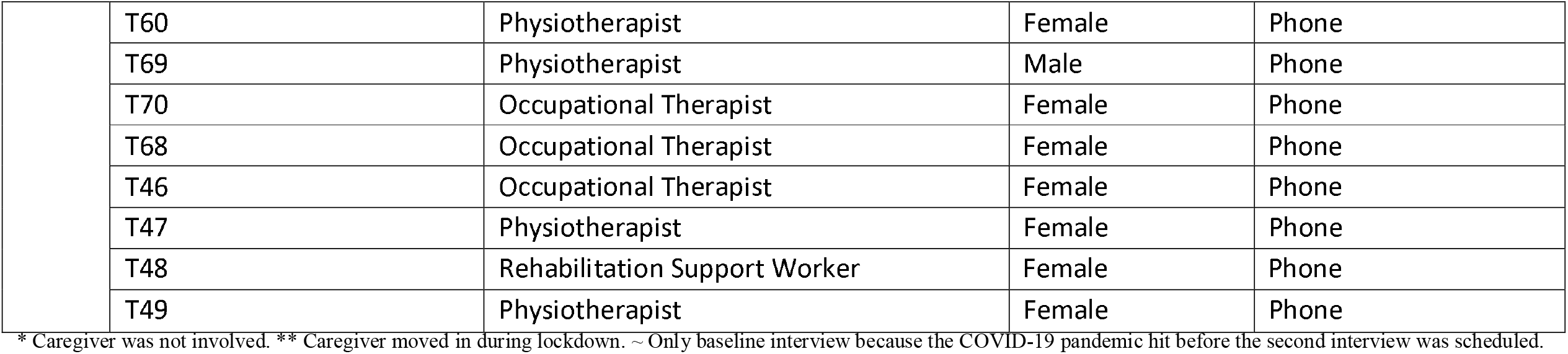
Participant-caregiver dyads and therapists interviewed in the study

| First set of interviews (i.e., pre-COVID-19) | Participant ID | Allocation group | Gender | Age range | Ethnicity | Living alone? | Caregiver ID | Age | Relationship to participant | Interview modality |
| --- | --- | --- | --- | --- | --- | --- | --- | --- | --- | --- |
|  | P1 | Control | M | 61-70 | White | Yes | C1 | 71-80 | Sibling | Dyadic face-to-face |
|  | P2 | Intervention | M | 81-90 | White | No | C2 | 81-90 | Partner | Dyadic face-to-face |
|  | P3 | Intervention | M | 81-90 | White | No | C3 | 61-70 | Partner | Dyadic face-to-face |
|  | P4 | Intervention | F | 71-80 | Black | No | C4 | 41-50 | Child | Dyadic face-to-face |
|  | P5 | Intervention | F | 81-90 | White | Yes | C5 | 51-60 | Child | Dyadic face-to-face |
|  | P6 | Control | F | 71-80 | White | No | C6 | 71-80 | Partner | Dyadic face-to-face |
|  | P7 | Control | M | 71-80 | White | No | C7 | 61-70 | Partner | Dyadic face-to-face |
|  | P8 | Control | M | 71-80 | White | No | C8 | 71-80 | Partner | Dyadic face-to-face |
|  | P9 | Control | M | 71-80 | White | No | C9 | 71-80 | Partner | Dyadic face-to-face |
|  | P10 | Control | F | 71-80 | White | No | C10 | 71-80 | Partner | Dyadic face-to-face |
|  | P11 | Intervention | F | 71-80 | White | No | C11 | 71-80 | Partner | Dyadic face-to-face |
|  | P12 | Intervention | M | 71-80 | White | No | C12 | 61-70 | Partner | Dyadic face-to-face |
|  | P13 | Intervention | M | 71-80 | White | No | C13 | 71-80 | Partner | Dyadic face-to-face |
|  | P14 | Intervention | M | 71-80 | White | No | C14 | 71-80 | Partner | Dyadic face-to-face |
|  | P15 | Control | M | 71-80 | White | No | C15 | 61-70 | Partner | Dyadic face-to-face |
|  | P16 | Intervention | M | 61-70 | White | No | C16 | 61-70 | Partner | Dyadic face-to-face |
|  | P17 | Intervention | M | 71-80 | White | No | C17 | 71-80 | Partner | Dyadic face-to-face |
|  | P18 | Intervention | M | 71-80 | White | No | C18 | 81-90 | Partner | Dyadic face-to-face |
|  | P19 | Intervention | M | 81-90 | White | No | C19 | 81-90 | Partner | Dyadic face-to-face |
|  | P20 | Control | M | 81-90 | White | No | C20 | 51-60 | Partner | Dyadic face-to-face |
| Second set of<br>interviews (i.e., during<br>COVID-19 pandemic) | P21 | Intervention | F | 91-100 | White | Yes | C21 | 61-70 | Child | Dyadic on phone |
|  | P22 | Intervention | M | 71-80 | White | No | C22 | 61-70 | Partner | Dyadic on phone |
|  | P23 | Control | F | 71-80 | White | No | C23 | 81-90 | Partner | Dyadic on phone |
|  | P24 | Intervention | F | 71-80 | Black | Yes |  |  |  | Individual* on phone |
|  | P25 | Intervention | M | 81-90 | White | Yes |  |  |  | Individual* on phone |
|  | P26 | Intervention | M | 71-80 | White | No | C26 | 71-80 | Partner | Dyadic on phone |
|  | P27 | Intervention | M | 71-80 | White | No | C27 | 81-90 | Partner | Dyadic on phone |
|  | P28 | Intervention | M | 81-90 | White | No | C28 | 71-80 | Partner | Dyadic on phone |
|  | P29 | Intervention | M | 91-100 | White | No | C29 | 71-80 | Partner | Dyadic on phone |
|  | P30 | Intervention | F | 71-80 | White | No | C30 | 71-80 | Partner | Dyadic on phone |
|  | P31 | Intervention | F | 61-70 | White | Yes |  |  |  | Individual* on phone |
|  | P32 | Intervention | F | 71-80 | White | No | C32 | 61-70 | Partner | Dyadic on webcam |
|  | P33 | Intervention | M | 81-90 | White | No | C33 | 71-80 | Partner | Dyadic on phone |
|  | P34 | Intervention | M | 71-80 | White | No | C34 | 71-80 | Partner | Dyadic on phone |
|  | P35 | Intervention | F | 71-80 | White | No** | C35 | 41-50 | Child | Dyadic on phone |
|  | P36 | Control | M | 71-80 | White | No | C36 | 71-80 | Partner | Dyadic on phone |
|  | P37 | Control | M | 71-80 | White | No | C37 | 71-80 | Partner | Dyadic on phone |
|  | P38 | Control | M | 81-90 | White | No | C38 | 81-90 | Partner | Dyadic on phone |
|  | P39 | Intervention | M | 71-80 | White | No | C39 | 71-80 | Partner | Dyadic on phone |
|  | P40 | Intervention | M | 71-80 | White | No | C40 | 51-60 | Partner | Dyadic on phone |
|  | P41 | Intervention | F | 91-100 | White | No | C41 | 71-80 | Partner | Dyadic on phone |
|  | P42 | Intervention | M | 71-80 | White | No | C42 | 81-90 | Partner | Dyadic on phone |
|  | P43 | Intervention | F | 81-90 | White | No |  |  |  | Individual* on phone |
|  | P44 | Intervention | F | 81-90 | White | Yes |  |  |  | Individual* on phone |
| First set of<br>interviews (i.e., pre-COVID-19) | Therapist ID |  |  | Profession |  |  |  | Gender |  | Type of interview |
|  | T45 |  |  | Occupational Therapist |  |  |  | Female |  | In-person |
|  | T46 |  |  | Occupational Therapist |  |  |  | Female |  | In-person |
|  | T47~ |  |  | Physiotherapist |  |  |  | Female |  | In-person |
|  | T48 |  |  | Rehabilitation Support Worker |  |  |  | Female |  | In-person |
|  | T49 |  |  | Physiotherapist |  |  |  | Female |  | In-person |

|  |  |  |  |  |
| --- | --- | --- | --- | --- |
|  | T50~ | Occupational Therapist | Female | In-person |
|  | T51 | Physiotherapist | Male | In-person |
|  | T52 | Rehabilitation Support Worker | Female | In-person |
|  | T53~ | Rehabilitation Support Worker | Female | In-person |
|  | T54~ | Rehabilitation Support Worker | Female | In-person |
|  | T55~ | Rehabilitation Support Worker | Female | In-person |
|  | T56 | Rehabilitation Support Worker | Female | In-person |
|  | T57 | Occupational Therapist | Female | In-person |
|  | T58 | Rehabilitation Support Worker | Female | In-person |
|  | T59 | Occupational Therapist | Female | In-person |
|  | T60 | Physiotherapist | Female | In-person |
|  | T61~ | Physiotherapist | Female | In-person |
|  | T62 | Occupational Therapist | Female | In-person |
|  | T63 | Rehabilitation Support Worker | Male | In-person |
|  | T64 | Physiotherapist | Female | In-person |
|  | T65 | Physiotherapist | Female | In-person |
|  | T66~ | Rehabilitation Support Worker | Female | In-person |
|  | T67~ | Rehabilitation Support Worker | Male | In-person |
|  | T68~ | Occupational Therapist | Female | In-person |
| Second set of interviews (i.e., during COVID-19) | T50 | Occupational Therapist | Female | Phone |
|  | T55 | Rehabilitation Support Worker | Female | Phone |
|  | T53 | Rehabilitation Support Worker | Female | Phone |
|  | T56 | Rehabilitation Support Worker | Female | Phone |
|  | T58 | Rehabilitation Support Worker | Female | Phone |
|  | T57 | Occupational Therapist | Female | Phone |
|  | T59 | Occupational Therapist | Female | Phone |
|  | T60 | Physiotherapist | Female | Phone |
|  | T69 | Physiotherapist | Male | Phone |
|  | T70 | Occupational Therapist | Female | Phone |
|  | T68 | Occupational Therapist | Female | Phone |
|  | T46 | Occupational Therapist | Female | Phone |
|  | T47 | Physiotherapist | Female | Phone |
|  | T48 | Rehabilitation Support Worker | Female | Phone |
|  | T49 | Physiotherapist | Female | Phone |

\* Caregiver was not involved. \*\* Caregiver moved in during lockdown. ~ Only baseline interview because the COVID-19 pandemic hit before the second interview was scheduled.

### General reception of the PrAISED programme

PrAISED was well received by participants and caregivers. Participants in the control group appreciated the brief falls assessment and advice provided by the team. While some reported increased awareness of exercise and health, this did not have any substantial impact on their physical activity levels pre-PrAISED:

*‘We are more aware of our health, so I do think it’s making a difference to our lifestyle, but not as much as it should, we would be better if we did more exercise’ C15*

*‘We were doing it before anyway Before PrAISED. We were all over the parks and Attenborough [nature reserve] and everywhere’ Participant’ P10*

When invited to give feedback on PrAISED during the interviews, 23/29 (80%) participants in the intervention group reported a very positive experience with PrAISED and that the programme had made a positive change in their lives. An example of the positive impact of PrAISED is illustrated by John’s story: https://vimeo.com/372693414/3f698b5f60. Participants and caregivers reported they valued therapist visits for the opportunity to improve or maintain physical ability and boost their confidence:

*‘It did make a difference, because at the start of the study I could only walk a short distance; whereas at the end of the study I could walk round the block, and it gave me quite a lot of confidence’ P12*

Therapy visits were an opportunity for participants and caregivers to develop awareness and knowledge about exercise and dementia, as well as to learn about opportunities for activities and resources available in the community:

*‘I think doing these things has enabled me now to think about it and challenge myself to improve my fitness and keep on doing things and hopefully whatever is the problem, it doesn’t develop into dementia’ P16*

*‘E. (therapist) who used to come, she told us about it (exercise class in the community). Apparently, it’s been going for about two years, but we didn’t know anything about it. So, he goes there every week on a Wednesday now for an hour. So that does him good’ C03*

Participants appreciated that therapists would arrange modifications to the house to make it more dementia-friendly:

*‘A. (therapist) arranged for him to have a perching stool, because he did try to do some potatoes one day. Well, he did so many and then he couldn’t stand up any longer. And E. (another therapist) fixed it to the right height and everything. I don’t think we can ask for anything better really’ C04*

The social interaction and support presented by therapy visits brought genuine affirmation and respect for the participants’ personhood. The interactions were described as ‘deep’ and ‘intimate’:

*‘It gave him something to look forward to. It was a little bit of structure to know that someone was coming every so often’ C12*

*‘I can say to the therapist if he’s not in a right mood today, I can talk to them, you know what I mean, and I’ll have a laugh’ C14*

*‘Being with R. (therapist) is very pleasant company, so that is good. I will miss his friendship and his ability to help me maintain what little memory I’ve got and the incentive that he gave me to continue physical exercise’ P18*

As a result of positive interactions and support from therapists, the participants were more willing/motivated to initiate and maintain exercise:

*‘When the support worker comes along, she encourages you to do them doesn’t she? You wouldn’t say no I’m not going to do them really’ P13*

### Facilitators to achieving positive intervention outcomes

The therapists were instrumental in supporting participants to achieve outcomes. Their ability to encourage, motivate and instil confidence to participants was highly regarded:

*‘She (PT) makes you feel as if you can do this, and this is a good thing to do and really gets you started and motivates you to start with’ C03*

Good rapport between therapists and participants/caregivers and the social element of therapy visits were key factors for positive outcomes. Feelings of loneliness or social exclusion of participants/caregivers were common, due to mobility issues, symptoms of dementia, demographic reasons (close friends passing away, caregiving responsibilities); therapy visits came to be seen as a change from the usual routine, an opportunity for human interaction at a meaningful level:

*‘It was mentally stimulating for him, and it was something to look forward to. It was someone to come and chat, it was someone different to me being in the house. And I think you get to befriend that person when she comes’ C12*

The therapy team provided a source of knowledge about dementia care, social care, pensions, tax exemption, medications, community groups, support, respite, and care planning. Participants and caregivers were often unaware of resources, activities, initiatives being offered in the community. The knowledge of the therapy team often bridged the gap between services and users. Receiving information about the potential benefits of the intervention was a key facilitator to ensuring participants’ commitment to PrAISED, as participants might have vague idea about the nature and purpose of the intervention:

*‘The participant could do with understanding the link between exercise and mood and feelings of wellbeing and potential improvements generally in cognition. That might harness motivation’. Therapist’ T57*

Caregivers were also facilitators to achieving positive outcomes. Given the support needs that dementia entails, relying on the caregiver to maintain exercise/activity levels in between therapy visits was common. Some caregivers were able and willing to provide support, as they anticipated the potential benefits of the programme, which extended to themselves and to their relationship with the person they cared for:

*‘I go with him because I don’t think he could cope with that. But by me going with him, I chat to other people that have got similar problems. And so, in a way I quite look forward to going because rather than the little circle I’ve already got, I see a little bit bigger circle’ C02*

Expectations and potential goals to be achieved as a result of taking part in PrAISED was linked to positive outcomes. For some, there was an expectation that the programme would improve mobility and independence:

*‘Hopefully I’m going to get better. My legs are going to strengthen. And I will be able to get up and walk about unaided, if you like’ P02*

Tailoring the programme and goals according to participants’ interests with a focus on achieving meaningful goals/purposeful activities (e.g., helping participants to continue doing the activities they enjoy doing) was a booster to some participants’ commitment to PrAISED and as a result positive outcome:

*‘I really just want to keep us as active as I possibly can. We’ve got great grandchildren, which I like to see as often as possible. I can’t pick them up and carry them about now’ P06*

Some participant-level variables were linked to positive outcomes. Experiencing benefits as a result of doing physical exercise promoted a sense of achievement. Some participants recognised benefits such as improvement in their physical health and coming to terms with dementia. Reaching goals that seemed impossible just months before boosted participants’ commitment to continue with the exercises, by progressing their intensity and setting new goals:

*‘I’m feeling more flexible now. I’m accepting it a bit more. I don’t get a sharp pain or anything. And if I keep moving and doing something I seem to get away with it all right’ P07*

### Barriers to achieving positive intervention outcomes

A prominent factor linked to negative intervention outcomes was pre-existing, often chronic physical health conditions. Participants’ physical health greatly affected delivery and response to PrAISED, as well as which benefits could be achieved following the intervention. Chronic health conditions, such as arthritis and its consequent pain, had a particularly negative effect on the ability to perform the physical activities set out in the programme. While some participants showed resilience by adopting compensatory strategies, such as concentrating physical activity on the days when they felt better, many reported either being unable to do the exercises and activities set out in their programme or being unmotivated to engage with no prospect of improvement:

*‘I was wondering whether I’d give up altogether because I’ve seen three different doctors at different times, and they can’t do anything with it (pain) and it just comes and goes. Well, if it’s going to do that it’s a bit pointless isn’t it, my trying to strengthen something that comes and goes’ P05*

Cognitive impairment and symptoms including apathy, forgetfulness, and fluctuations in wellbeing posed a particular barrier to success. A common challenge was that memory impairment would cause the participants to forget they were committed to the exercise programme. Thus, the therapists found it difficult to progress participants toward final intervention goals:

*‘I don’t know if when I arrive, she’s (participant) going to be like “R. lovely, let’s do the exercises”, and be really on the ball, or I could go in, she could be quite anxious about me being there because she needs to go and pick up the kids from school. And so that’s why I can’t plan a session’ T58*

A major issue experienced in PrAISED was around tapering therapists’ support over time. Participants contended that habit formation was challenging given the support needs of people living with dementia, and that reducing the number of visits over time was inevitably linked to a reduction in the amount of physical exercise and obtainment of associated benefits:

*‘R. (therapist) came twice a week, once a week and then once a fortnight. And then all of a sudden it was once a month. And when it went down to once a month it didn’t keep me on my toes to religiously do the exercises’ P16*

This was particularly challenging for participants who lived alone who could not count on the support or reminders of caregivers outside therapy sessions. Some personal characteristics of the participant also presented barriers. While a sense of competitiveness could push some participants to aspire to achieving more from the programme, many tended to compare their present self to past achievements. This generated a sense of defeat, which was hard to challenge. This was particularly visible in male participants who had a previous career that emphasised physical ability and performance excellence:

*‘The fact that I used to swim 50 lengths every time I went and now the thought of being able to do five or six isn’t really a motivation’ P03*

Another barrier was unrealistic or unachievable expectations of some participants, which would set them up for failure. Unrealistic expectations might have been generated by wishful thinking about “curing” dementia or miscommunication with the therapists:

*‘The thing is mum had been told that she could go with her scooter on the bus, and the truth is you can’t. The difficulty with that is they give you these ideas that it’s all possible, and realistically it’s not’ C05*

The realisation that the unachievable goal had not been obtained would inevitably affect participant’s adherence to the intervention and led to not achieving positive outcomes. Some caregiver factors presented barriers, such as their views on dementia and attitudes toward risk. Some caregivers feared that the person they cared for might be at risk of falling or harm when doing PrAISED exercises or activities in between therapy visits. The anxiety was particularly acute where there was a history of previous falls and injuries. They would become instinctively risk-averse and potential gatekeepers to the participant levels of activity:

*‘I’ve seen what happens when he falls, and I’ve got that awful fear. And all I think about is oh my god he’s going to fall, he’d going to fall. I think I couldn’t walk him round the block, I’d be an absolute, well I would, I’d be shaking by the time I got back’ C12*

Given the support needs of participants to be able to maintain exercise/activity in between therapy sessions, carer input (and care responsibilities) would inevitably increase as a result of PrAISED. Some caregivers felt that the added burden on top of existing caring duties was too much, and they could not fully support the participant:

*‘A participant, it was his wife who said to me I can’t support my husband to engage in the programme as much as I wanted to. It’s causing me stress, too much pressure, I’m going to have to withdraw. So, it wasn’t actually the participant, but it was his wife’ T57*

Several practical and logistical factors posed barriers to a successful intervention. Life often ‘got in the way’ of doing the exercise programme. The participants and caregivers often found it difficult to find time to be allocated to the programme, caught as they were in their daily routines:

*‘We actually are very poor at keeping up with that because your life is taken over by your normal routine. Our normal routine is actually quite busy’ C15*

Living alone posed a barrier as well, because of the lack of reminders to initiate and support to facilitate exercise/activity. Accessibility of home for exercise or activity, and lack of opportunities in the community was a factor. Some participants reported they struggled to achieve goals that involved outdoor activities, given risks and accessibility. This was a particular concern for people with dementia living alone or in rural areas:

*‘I cycle in the gym, but I don’t go anywhere. I find these roads quite frightening. There are so many potholes, the traffic is very big here, in the summertime lorries, tractors, great big farm implements’ P15*

Unexpected events beyond the control of the therapy team posed barriers to the intervention, including injuries, hospitalisations, medications, holidays, and other life events, as the progress made could easily be halted or lost:

*‘That (new medications’ side effects) altered the programme as well because we had to get over that. And I think since then you’ve sort of lost’ C12*

A unique circumstance that disrupted the intervention was the COVID-19 pandemic. Some participants were negatively affected by the lack of face-to-face support and lost the progress they had previously made. Remote delivery lacked the human connection that had been instrumental to the success of PrAISED pre-COVID-19 pandemic:

*‘From Mum’s point of view, not used to using these types of technologies it’s not just like having someone sat in the armchair next to you having a chat. It’s just not what she is used to’ C21*

Remote support presented specific barriers relating to cognitive impairment. For example, not seeing a face during telephone sessions prevented participants from recalling who they were talking to. Remote delivery of PrAISED proved challenging for caregivers too, who experienced an increase in their support role in the lack of in-home visits from the therapists. Their respite time reduced, as caregivers reported needing to do the exercise routine with participants:

*‘I think I would prefer her (therapist) to come because I think when she was here, I could go into another room if necessary and do a job or two’ C28*

### Ecological system of PrAISED

It is important to emphasise that PrAISED was a complex intervention, and facilitators and barriers were generated through a complex interaction. Depending on the individual participant and the context, some facilitating mechanisms could also become barriers to intervention outcomes. For example, good rapport with therapist could be productive or counterproductive. Some participants might develop a habit of physical exercise though good rapport with the therapists that would be maintained post-PrAISED, while others might develop overdependency and give up exercise once therapist support was discontinued.

Some facilitators/barriers were dependent on PrAISED and modifiable (e.g., therapist’s support), while some others such as participants’ history, and unexpected events were not preventable or modifiable by the study team. However, they inevitably affected intervention outcomes.

No single facilitator or barrier was at play in equal measures in different participants. Different combinations/dosage of facilitators/barriers produced different outcomes (e.g., pain + loss of confidence vs. pain + loss of confidence + risk-aversion). They added or detracted from each other within a complex ecological system. The model in Figure 1 illustrates this complex interaction. As a result of this complexity, each participant had a distinct experience of the intervention. Some participants had a very positive response and experience to PrAISED, while others less so. The case study vignettes (Appendix 6) were developed to reflect the diversity of experiences of PrAISED.

## Discussion

The PrAISED RCT found that after 12 months, the Disability Assessment for Dementia (DAD) [21], and a range of other health status measures including balance, activity and quality of life were no different between intervention and control groups [13]. The aims of the process evaluation were to investigate the reception of the programme and explain findings from the RCT by identifying mechanisms, facilitators and barriers to achieving positive outcomes.

The PrAISED intervention was liked by participants with dementia and their family caregivers. We determined that several aspects of the strategy to promote engagement and motivation were successful, including delivery at home, by expert healthcare professional staff, goal setting, tailoring according to interests, co-morbidities and abilities, a focus on achieving practical and useful activities, and close involvement of family carers. Family and other carers were supportive and helpful, despite their own experience of strain and ill health, and their own ‘respite’ time being diminished. Positive risk taking was met with some scepticism, but efforts to increase confidence and planning to minimise risk were successful.

We included participants with relatively mild cognitive impairment, but forgetfulness and apathy proved to be major barriers to undertaking activities without direct supervision, carry-over between sessions and subsequently progression. This was a particular problem for people living alone. Co-morbidities, illnesses and injuries, and other disruptions were frequent and set back functional gains. Maintaining adherence to the PrAISED exercise and activity programme required active management. Habit formation was found to be at odds with the memory problems, and apathy typical of dementia [22]. Therapists reported a particular challenge linked to memory problems, whereby motivation for the intervention had to be developed anew during each session. This would make setting and achieving goals difficult, reduce margin for progress, and had a direct impact on outcomes. An important implication for future practice is that it is unlikely that an intervention can be successful with this population unless there is a recognition that support from significant others or therapy teams [23-26] is a pre-requisite for success. This also highlights the issue of maintenance of physical activity in those who live alone or lack support.

The COVID-19 pandemic and its consequent lockdown occurred mid-RCT. Access to community facilities and opportunities to keep active were curtailed. PrAISED therapy sessions needed to be adapted from face-to-face in-home to remote delivery through telephone or videoconferencing for a six-month period. Challenges linked to effective remote delivery included logistical factors, such as information technology accessibility and use. In line with our previous findings [27,28], this study found that remote support for some people living with dementia was feasible, but required pre-conditions that were often lacking, such as proper infrastructure (e.g., an internet connection), and support to use technology. This may be less of an issue in the future when older people will have used IT throughout their lives, but currently skills need developing and supporting to ensure that interventions requiring remote or hybrid (face-to-face/remote) delivery (e.g., due to remote location or mobility issues) [29,30] are equitably accessible to people living with dementia.

Despite the RCT results, participant responses to the intervention were positive. Participants valued the intervention as proactively addressing health issues that were of concern to them. Insofar as the intervention was successful, it was in ways that were not specifically intended or anticipated, including developing therapeutic relationships, affirmation of personhood, agency and hope, social contact and occupation, information giving and advice. The PrAISED RCT may not have identified outcomes that really mattered to participants and as a result did not capture them.

Benefits in the areas of social contact, interaction, information, and advice were gained largely due to therapists’ skills. Rehabilitation work with people living with dementia is complex and challenging, requiring specialist knowledge and skills to address the complex and distinct needs of this population [31]. Alongside therapists’ professional assets, the participants and caregivers appreciated a set of “soft” skills intrinsic to individual therapists that were instrumental to enhance intervention experience and engagement, including empathy, positivity, humaneness, active listening, and showing commitment to the programme/participants. Initially, good rapport was intended as a tool to maximise intervention uptake. In time (partly due to the social restrictions imposed by COVID-19), participants came to consider therapy visits as a means for social interaction with the therapist, and advice, something that they greatly valued. Often, the emphasis came to be placed on the social occasion of the therapy visits, rather than on exercise. The implication for future interventions is to acknowledge that social exclusion is common among people living with dementia (and caregivers) in certain conditions (e.g., living alone, living remotely) and that integrating intervention protocols with strategies fulfilling participants’ needs for meaningful intimate human connectedness will boost their experience of the intervention. Such strategies could include, as in the PrAISED example, therapists linking participants to opportunities for social inclusion in the community (and measures to detect greater or less social inclusion).

Another important precondition for intervention engagement was caregiver support. An example of the impact of caregiver support was embracing positive risk-taking, which could encourage participants’ activity levels. An effective way to ensure caregiver support was to address (justified or unjustified) concerns, resistance, and pre-conceptions about physical activities in dementia. Another way to garner caregiver endorsement was to build their capability to provide support to the person, as it was recognised that this would increase care burden. Therefore, for any future successful intervention programme, caregivers’ emotional, physical, and financial burden [32-44] should be acknowledged and effectively addressed through a holistic approach, where the caregiver is also considered as the recipient of a package of support/care.

## Strengths and limitations

This study presents novel data and implications for research, clinical practice and a framework for future process evaluations. The extraordinary circumstances of the COVID-19 pandemic presented problems for both intervention delivery and research, but resulted in novel data [45,46]. We used an innovative model of Patient and Public Involvement co-research [47].

The evaluating team was mostly but not fully independent of the intervention delivery team. Risk of bias was minimised through the involvement of multiple coders in the analysis of the interviews external to the delivery team. Data generated through the interviews might not reflect the experience of all participants in PrAISED given the relatively small sample. The process evaluation adopted purposive sampling to ensure representation of different experiences. In dyadic interviews, some caregivers were reserved in discussing sensitive subjects in the presence of the person they cared for. Whilst recognising this limitation, the team also believed that existing dynamics between caregiver and participant did not need addressing, as they represented the bedrock (i.e., ecological system) on which the intervention was delivered.

## Conclusion

We conclude that either the content of the intervention was ineffective, or that progression and symptoms of dementia or intercurrent health crises overwhelmed any beneficial effects. Our intervention was ambitious, and at the limit of what a public healthcare service might provide in terms of intensity. We identified factors that can successfully support engagement in interventions. It is possible at a more intensive intervention might have had beneficial effects, but the goal of maintaining or slowing decline in independence and activity in dementia may be impossible. This raises the question of whether subjective wellbeing and health gain in dementia is better achieved through socio-emotional-relational interventions rather than through bio-medical remediation. Maintenance of functional ability is valued, but in the face of inevitable progression of disease, other less tangible outcomes become important, challenging how we frame ‘health gain’ and trial outcomes.

## Data Availability

All data produced in the present study are available upon reasonable request to the authors

## Acknowledgments

The PrAISED team would like to thank Katarzyna Kowalewska for her invaluable support in this study.

## Appendix 1. Interviews conducted as part of the study

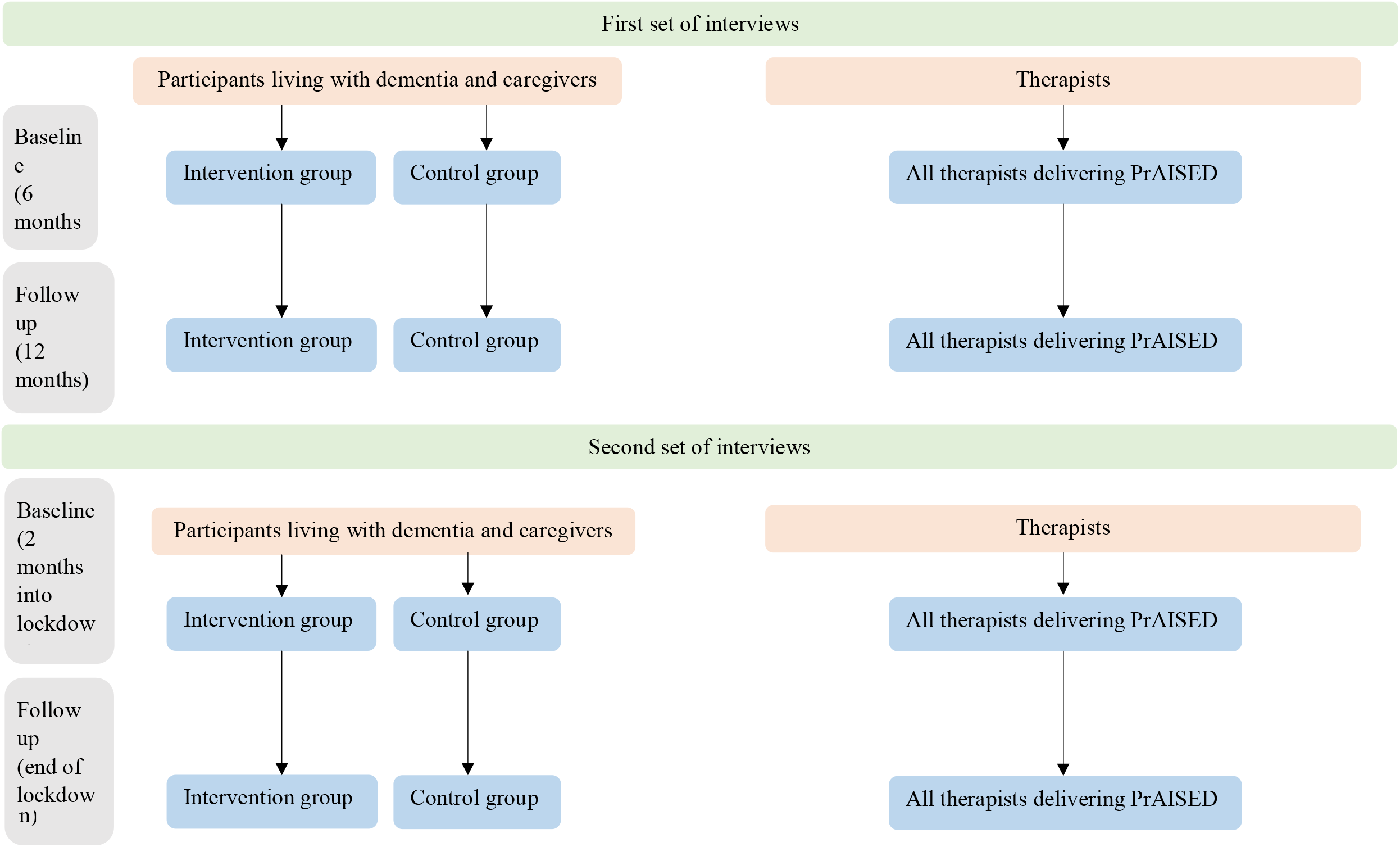

## Appendix 2. Topic guide for participants with dementia and caregivers (First set of interviews)

### Pre-interview

- Researcher introduces himself and engages in small talk to break the ice with participants (e.g., give thanks for being invited over, gives compliments about the home, and asks how the person is doing on the day).
- Researcher explains his professional role and the purpose of the visit
- Researcher goes through the Information Sheet with the participants. The following will be clearly explained:
  1. The interview will be audio-recorded to have an accurate record of what was said
  2. Anything mentioned during the interview is confidential and no one except members of the PrAISED research team will know what was said
  3. In using any information in a report, the participant’s anonymity will be maintained
  4. Participation is totally voluntary
  5. The participants can withdraw at any time and the research team can use the information collected thus far, unless the participants specifically withdraw consent for this.
- Researcher asks if participants have any concerns / questions / doubts.
- Researcher seeks informed consent
- Researcher asks participants if they are comfortable being interviewed together or they prefer being interviewed alone

### Interview

#### General questions

1. Do you feel that being involved in the study has been beneficial?
2. If so, what are the positive results of the activities?
3. Have you experienced any negative effects of the activities?
4. Do you think the programme has enabled you to enjoy more your daily activities?
5. I would like to start by asking your views around exercise…

#### Personal beliefs

- How important do you think being active is to help people stay healthy?
- How important do you think being active is to help people stay independent?

#### Motivation

- Why did you decide to take part in the programme?
- Were you encouraged by anyone to take part or was it your own choice?
- What helps you keep going with the programme?
- On a scale from 1 to 10, how much do you feel you want to continue with the Activities, once the programme has finished?

#### Autonomy and control

- Is it important for you to decide what you do, or do you prefer to leave it to others?
- (If yes to previous question), how much have you been able to make those decisions?
- How could we make you feel more involved?

#### Intervention characteristics

- Does the programme of physical activities suit your needs and preferences?
- What part of the programme of physical activities do you like the most?
- What part do you like the least and how could this be improved?

#### Self-efficacy and emotional support

- Do you feel you are able to do the activities as well as you would expect?
- Do you have any concerns or anxiety about taking part in the programme/doing the activities?
- Did you receive encouragement and support from your therapist(s) and caregiver(s)?
- Is there anything that would help you feel more confident to do the activities?

#### Support (Practical)

- Do the therapists give you practical support? For example, do they show you how to do the activities, when to do them and where to do them?
- Does your (caregiver role) give you practical support? For example, does he / she remind you how to do the activities, when to do them and where to do them?
- What could be done to better support you?

#### Independence

- How has the study programme affected you? (e.g., on your health and activity)
- Has it given you greater independence?
- Have you noticed a change in your quality of life?
- Are there any activities you would like to be able to do that are not part of the programme?

#### Expectations

- Have you any personal goals you would like to achieve from the study?
- If yes, what goals are you looking to gain?
- Do you think you can achieve these goals, and do you need support to do this?

### Final remarks

- Any final thoughts and feedback on the programme?
- Would you be happy to meet up again in three months’ time to see how you are doing?

## Appendix 3. Topic guide for therapists (First set of interviews)

### Pre-interview

My name is ………………………., I am a ………………………. working on the PrAISED trial.

This interview is for the PrAISED process evaluation; for this we are aiming to carry out interviews with participants, caregivers and therapists, to look at how the intervention works.

The session is going to be digitally audio-recorded, it is totally confidential, participation in the interview is voluntary and you can withdraw at any time. Do you give your consent to take part?

Can I ask you to introduce yourself and what your professional experience of working with people with dementia is?

Do you have any questions or concerns about the interview?

### Instructions

- *Before asking the questions explain each heading before asking the questions i*.*e. ‘I would like to ask you some questions about your motivation*
- *You may have to pick a couple of questions from each section if the interview is taking a long time (this could be varied for each participant*
- *Please ask all questions in red*

### Interview

#### General questions

1. What is your understanding of PrAISED?
2. Can you explain your experiences of the programme so far?
3. What are your views on the programme’s effectiveness?
4. I would like to start by asking around your views on physical activity *(if interviewee is physiotherapis*t), on activities of daily living *(if interviewee is Occupational Therapist or support worker)*…

#### Personal Beliefs

5. Do you feel that your views have altered since taking part in PrAISED?

#### Motivation

6. Why did you become involved in the programme?
7. What aspects of the training and delivery of PrAISED have a positive or negative impact on your motivation?
  a. Prompt re negative or positive if needed
8. Is there anything that could be done to increase/maintain your motivation to continue using the programme?

#### Expectations

9. What expectations do you have of your role within PrAISED?
  a. i.e., what were your expectations of being part of the PrAISED project?
10. Have your expectations that you had at the beginning of PrAISED been fulfilled?
11. What are your professional goals in PrAISED?
12. Did you anticipate any barriers to delivering the programme?
  i. Can you explain these?
13. Did you anticipate any facilitators to delivering the programme?
  a. i.e., what did you think would help you deliver the intervention?
    i. Can you explain these?
14. Did you anticipate that the intervention would improve the quality of life of the participants and their caregivers?

#### Autonomy and control

15. Did you have as much input as you would like in tailoring PrAISED for individual patients?
16. To what extent do you feel that your input as an experienced therapist is valued by patients / caregivers?

#### Self-efficacy

17. How competent in your professional role do you feel, to deliver the intervention?
18. What could be improved in the training to boost your confidence to deliver the intervention?

#### Support (Practical and emotional)

19. What support have you had to take part in the PrAISED programme
  a. How supportive is your PrAISED clinical team?
  b. How supportive are your colleagues outside of PrAISED?
20. How useful has contact with the University been?
  a. what in particular has been helpful?
21. How have you found the training you received in PrAISED (e.g., initial training and ongoing support, like the teleconferences)?
  i. What in particular was successful or not successful (Case studies, discussions?
22. Did you find the training met your needs in the way that you like to learn?
  i. Can you explain this
23. Did you feel you had enough training to effectively deliver the intervention?
24. What could be done to make you feel better supported whilst involved in PrAISED?
25. How collaborative do you feel that the person with dementia and their caregivers are in undertaking the PrAISED programme?

#### Intervention characteristics

26. How much do you feel that the intervention fits into your aspirations and professional development as a therapist?
27. Are there any aspects of the programme think work effectively or don’t work effectively?
  a. Prompt re. work effectively/don’t work effectively
28. How could the programme be improved?
29. How does your involvement in PrAISED fit into your working routine?
  a. Do you feel overburdened as a result of taking part?

#### Final remarks

30. Any final thoughts on the programme?
31. Would you be happy to meet up again in three months’ time for further feedback.

## Appendix 4. Topic guide for participants with dementia and caregivers (Second set of interviews – i.e., during the COVID-19 lockdown)

### Pre-interview

- Researcher introduces himself and engages in small talk to break the ice with participant (e.g., give thanks for accepting to talk over the phone, asks how the person is doing on the day).
- Researcher explains his professional role and the purpose of the call
- Researcher ensures that the participants have read the Information Sheet (previously sent through mail), prior to signing the consent form. The following will be clearly explained:
  1. The interview will be audio-recorded to have an accurate record of what was said
  2. Anything mentioned during the interview is confidential and no one except members of the PrAISED research team will know what was said. The only circumstance where confidentiality will be breached is if during the interview, information is disclosed about a potential risk of harm for the participant. In this case, the information will be reported by the researcher to a senior clinician within the PrAISED team and an action plan discussed and implemented, as appropriate.
  3. In using any information in a report, the participant’s anonymity will be maintained
  4. Participation is totally voluntary
  5. The participant can withdraw at any time and the research team can use the information collected thus far, unless the participant specifically withdraws consent for this.
- Researcher asks if participant has any concerns / questions / doubts.

### Interview

Just to explain, I would like to get your opinion on the effect the recent coronavirus has had on you, but first want to ensure that you are happy to discuss this. It has been a difficult time for everyone, and I would not want to cause any extra stress. Are you comfortable talking about the impact the recent changes in PrAISED caused by the coronavirus pandemic have had on you?

(Continue if yes)

Thank you, I would like to start by asking what impact the recent changes in PrAISED due to the coronavirus pandemic have had on you…

As an introductory question, have you stayed in the house? Has this made you feel more isolated and lonelier?

#### Personal beliefs

How important do you think being active is now that you are staying at home?

Have you been thinking more about your health now than you were before coronavirus? Could you tell me more?

#### Motivation

Are you able to carry on being active while at home?

Is there anything that helps you keep going with the programme, now that you are at home?

How much do you want to continue with the activities, now that you are at home?

Is there anything else that would help you keep active whilst you are unable to go out?

#### Autonomy and control

How do you feel now that you need to stay at home and cannot do the activities you like outdoors? Do you feel less in control of your daily activities?

#### Intervention characteristics

Have you been able to follow the PrAISED programme as before?

How have you felt about receiving the therapists support by telephone (substitute with any other means used)?

What have been the positive and negative changes with this new approach?

Do you think the PrAISED programme is as effectively delivered without face-to-face interaction? If yes, have you any thoughts on what characteristics would make it work better for you?

#### Self-efficacy

Do you feel you are still able to do the exercises and activities now that you therapists cannot visit you in person, due to the Coronavirus pandemic?

Have you still received encouragement and support from your therapist(s) and caregiver(s)?

Is there anything that would help you feel more confident to do the activities?

#### Social opportunity and emotional support

How did it feel when your therapist could no longer visit you?

How has staying at home changed your social life? Are you able to talk to other people outside your home?

Have you found other ways to socialise with others (phone, computer)?

Is there anything that would have helped you feel more emotionally supported?

#### Support (Practical)

Do the phone sessions (substitute with any other means used) that you receive from therapists help you understand how to do the activities, when to do them and where to do them?

Have you experienced any problems trying to do the exercises without the therapists being with you?

Have you been more worried about falls without face-to-face support from the therapists?

Does your (caregiver role) give you practical support? For example, does he / she remind you how to do the activities, when to do them and where to do them?

Does your (caregiver role) give you more support, now that the therapists are not visiting? What could be done to better support you to do the exercises?

#### Independence

How has staying at home affected your independence? (e.g., on your health and activity)

Do you feel more dependent on others?

Have you noticed a change in your quality of life?

Are there any activities you would like to do that you cannot do at home?

#### Expectations

Have your personal goals changed as a result of staying at home due to the coronavirus?

If yes, what goals are you looking to gain now?

Do you think PrAISED is supporting you to achieve them?

### Final remarks

Any final thoughts and feedback on the programme?

## Appendix 5. Topic guide for therapists (Second set of interviews – i.e., during the COVID-19 lockdown)

### Pre-interview

My name is ………………………., I am a ………………………. working on the PrAISED trial.

Do you have any questions or concerns about the interview?

### Interview

To begin with, what is your overall view around the impact of the change from face-to-face support to remote support in PrAISED?

What do you feel the main barriers to the remote support have been, for you as a therapist and for participants?

Have you been able to apply the PrAISED principles remotely? How has your clinical practice changed?

Have you been able to use any video support, or did you only work on the phone? How did you find this shift?

How do you feel that the participants have responded to the shift from face-to-face to remote support?

How do you feel in relation to the support that you have been given by your team and the PrAISED team?

How do you feel that participants and caregivers have responded in terms of motivation to keep engaged in the process during lockdown?

How do you feel in a relation to video support? Is it something implementable, is it something that is out of the question with this population?

Do you feel that overall, in this lockdown situation the caregivers have become given more central in supporting the participants (and therapists) to engage with PrAISED? How has the relationship and dynamic between participant, caregiver and therapist changed as a result of remote delivery?

Looking back at these last 3 months, do you think there has been any value in doing this? Have there been any unexpected positive in this new way of delivering PrAISED?

What is your view now on PrAISED, what are your expectations? Have they changed? What has PrAISED become for you now?

What can we learn from the experience of the lockdown for future implementation of interventions? Do you have any new insight that might be helpful for the future?

### Final remarks

Any final thoughts and feedback on the programme?

## Appendix 6. Case-studies

### Case study 1. Interaction of factors generating a negative experience of PrAISED

P1 is a man in his early 80s; C1 his caregiver in her 60s. The participant and the caregiver had very different views on PrAISED and the participant only participated in the programme through the caregiver’s insistence. While the participant was compliant throughout the programme, he only engaged passively without a commitment to achieving goals. Although the participant recognised that his physical health had deteriorated over time, he seemed quite contented with where he was and with his sedentary activities, including reading newspaper and doing sudoku. The caregiver was of the opposite view and showed frustration at the participant’s attitude. The caregiver also felt that the participant’s deterioration was the cause of her staying at home most of the time and becoming socially isolated. She was much younger than the participant and she contended that she aspired to have a more active life outside the house. The caregiver appeared depressed and unable to create a constructive communication with the participant. She reported constantly trying to encourage the participant to engage with PrAISED and/or to do physical activity (e.g., walking together), but to no avail. As well as contending that he was satisfied with his life, the participant reported a lack of motivation to engage in exercises that were seen as too simple, repetitive, and boring. Further, engagement was hampered by a sense of defeat, due to the constant comparison with his former self (he had been a PE teacher and professional swimmer) and the awareness that he would never be able to match his previous physical fitness. He was therefore reluctant to do his swimming sessions out of the inevitable disappointment in himself. This led to no achievement of benefits from PrAISED, bar the social opportunities presented by therapy visits, which inevitably finished at the end of the 12-month programme.

### Case study 2. Interaction of factors generating a positive experience of PrAISED

Participant 7 is a man, Caregiver 7 is his partner, both in their late 70s. In the first interview, the participant and caregiver reported that the participant was resistant to accept the inevitable changes that ageing, and dementia entailed. The participant had been a very active cyclist in the past and was struggling to accept his deteriorating physical health. In the interview, the word dementia was only rarely used, and the participant kept hinting at the fact that the changes he was experiencing were typical of ageing. The caregiver downplayed the symptoms of dementia in an evident attempt to safeguard the person’s emotional health and self-esteem. The participant reported feeling depressed. He had experienced several falls and had lost confidence in himself. At the time of the first interview, the participant wished to be able to walk independently again to his local bowls club, which was the centre of his social life. To achieve this, he appeared very committed to the programme and had the full support of his partner and his family. The caregiver, however, appeared also extremely anxious about his partner’s risk of falling, which made her risk averse. At the time of the interview, the couple were still re-negotiating social / domestic roles (what to do, what is dangerous). In the second interview, both participant and caregiver reported experiencing great progress. Through full engagement with the programme, no more falls had occurred. The participant had been able to get fitter and regain his balance and confidence. There had been a boost in his quality of life, because he had been able to accomplish his goal (walking to the bowls club and spending time with his friends). He confirmed he felt emotionally better and accepting the changes of dementia, though he was still struggling to fully come to terms with the condition. The caregiver appeared less anxious and more positive about the effects of PrAISED as well. Both participant and caregiver were positive that the participant would continue to exercise after the end of PrAISED.

### Case studies 3 and 4. Same factor (rapport with therapist) being a facilitator in one participant and a barrier in another

For participant P11, the good rapport developed over time with the RSW proved a key ingredient in ensuring uptake of the exercise programme and the benefits associated with it. P11 had a history of being a very independent woman, who had been active and into sports throughout her life. She had a strong support system in place through her partner’s commitment to keep her active. This was further compounded by access to private personal training who boosted her opportunities to keep moving. In the presence of all these facilitators, good rapport with and support from the RSW functioned as a further enabler of the participant’s independence. P18 on the contrary reported having no friends and that he had minimal social contact. He lived with his partner in a rural area, far from opportunities to exercise in the community. Country roads were not conducive to independent walks as the associated risks were aplenty. In this case, the RSW came to be seen as a unique (and the sole) opportunity for exercise, sociality, and a change in routine. P18 developed feelings of attachment and dependency to the RSW and close to termination of support feelings of anxiety had developed. The participant stated that he would “deteriorate if he (the RSW) isn’t looking after me” and that without him he “couldn’t cope”. This resulted in the participant being pessimistic about the future and whether he would be able to continue his exercise routine.

